# Environmental factors and mobility predict COVID-19 seasonality

**DOI:** 10.1101/2021.09.15.21263648

**Authors:** Martijn J. Hoogeveen, Aloys C.M. Kroes, Ellen K. Hoogeveen

## Abstract

**Background:** We recently showed that seasonal patterns of COVID-19 incidence and Influenza-Like Illnesses incidence are highly similar, in a country in the temperate climate zone, such as the Netherlands (latitude: 52^°^N). We hypothesize that in The Netherlands the same environmental factors and mobility trends that are associated with the seasonality of flu-like illnesses are predictors of COVID-19 seasonality as well.

**Methods:** We used meteorological, pollen/hay fever and mobility data from the Netherlands with its 17.4 million inhabitants. For the reproduction number of COVID-19 (R_t_), we used data from the Dutch State Institute for Public Health. This R_t_ metric is a daily estimate that is based on positive COVID-19 tests in the Netherlands in hospitals and municipalities. For all datasets we selected the overlapping period of COVID-19 and the first allergy season: from February 17, 2020 till September 21, 2020 (total number of measurements: n = 218), the end of pollen season. Backward stepwise multiple linear regression was used to develop an environmental prediction model of the R_t_ of COVID-19. Next, we studied whether adding mobility trends to an environmental model improved the predictive power.

**Results:** By means of stepwise backward multiple linear regression four highly significant (p value < 0.01) predictive factors are selected in our combined model: temperature, solar radiation, hay fever incidence, and mobility to indoor recreation locations. Our combined model explains 87.5% of the variance of R_t_ of COVID-19 and has a good and highly significant fit: F(4, 213) = 374.2, p-value < 0.00001. The combined model had a better overall predictive performance compared to a solely environmental model, which still explains 77.3% of the variance of R_t_, and a good and highly significant fit: F_(4, 213)_ = 181.3, p < 0.00001.

**Conclusions:** We conclude that the combined mobility and environmental model can adequately predict the seasonality of COVID-19 in a country with a temperate climate like the Netherlands. In this model higher solar radiation, higher temperature and hay fever are related to lower COVID-19 reproduction, and mobility to indoor recreation locations with increased COVID-19 spread.

**Highlights:**

- The seasonality of COVID-19 can be well-explained by environmental factors and mobility.
- A combined model explains 87.5% of the variance of the reproduction number of COVID-19
- Inhibitors of the reproduction number of COVID-19 are higher solar radiation, and seasonal allergens/allergies.
- Mobility, especially to indoor recreation locations, increases the reproduction number of COVID-19.
- Temperature has no direct effect on the reproduction number of COVID-19, but affects mobility and seasonal allergens.
- Adding mobility trends to an environmental model improves the predictive value regarding the reproduction number of COVID-19.

## 1. Introduction

COVID-19 appears to be subject to multi-wave seasonality [1, 2], comparable to other respiratory viral infections and pandemics since time immemorial [3, 4]. It is observed that the COVID-19 community outbreaks have a pattern that is similar to those of other seasonal respiratory viruses [5, 6, 7, 8], whereby the seasonal dips coincide with allergy season in regions in the temperate climate zone [9, 10, 11]. The same factors that drive the seasonality of flu-like illnesses, appear to drive COVID-19 seasonality: solar radiation including ultraviolet (UV) light, temperature, relative or specific humidity, seasonal allergens (pollens) and allergies, and behavior. Regarding behavior, mobility data show the beneficial effect of restrictive measures on the effective reproduction number (R_t_) of COVID-19 [12, 13, 14], but the seasonal aspects of mobility are often overlooked. For example, during nice weather people spend more time outdoors. For flu-like illnesses, we previously showed that a compound predictor of solar radiation and seasonal allergens is highly significant though moderately strong r(222) = ‒0.48 (p < 0.001) [9]. It is unclear why environmental factors, such as higher solar radiation, a higher level of seasonal allergens (pollens) and subsequently hay fever are consistently associated with a lower R_t_ of COVID-19, and, thus possibly associated to COVID-19 seasonality as well. Exposure to solar radiation might be associated with better COVID-19 outcomes [15], and daylight is understood to regulate melatonin levels, and subsequently circadian (lung) immunity [16]. Further, increased UV light levels are associated with a more rapid degradation of SARS-CoV-2 particles [17], although the clinical relevance of this effect is debatable.

Upon the observation that allergic diseases are associated with lower rates of COVID-19 hospitalizations [18, 19], several pathophysiological explanations are provided, such as a lower expression of membrane-bound angiotensin-converting enzyme 2 (ACE-2) [20, 21], higher eosinophil counts [22, 23, 24], a reduced risk of a cytokine storm and hyper-inflammation [25], and T cell-mediated immune responses to allergens which might be effective against COVID-19 as well [26]. On the other hand, a recent international epidemiological study reported a positive correlation between pollen concentrations and COVID-19 incidence [27]. As another study, from Spain, could not confirm the latter finding [28], this is still a matter of considerable debate.

Further, we noticed that an estimate of R_t_ discriminates better between independent variables than incidence metrics [9], as it appears to be a more volatile or sensitive metric, includes incubation time lags, and is corrected for test bias and independent of seasonality. The reproduction number has also become the standard in predictive modelling for COVID-19..

Our hypothesis is that a model, combining both environmental factors and mobility trends, improves the prediction of the seasonality of COVID-19 compared to each factor alone. Therefore, the main objective of this study is to explore a model, including both environmental factors and mobility trends of people, to improve the prediction of the reproduction number for COVID-19 during spring season which coincides with the low-season of flu-like respiratory diseases in a country in the temperate climate zone such as the Netherlands (latitude: 52°N).

## 2. Methods

### 2.1 Data

#### 2.1.1 Reproduction Number for COVID-19

For the observations of R_t_, we used the respective dataset from the Dutch State Institute for Public Health (Rijksinstitutuut voor Volksgezondheid en Milieu; RIVM) [29] from February 17, 2020 till September 21, 2020. RIVM uses a standard method to calculate the R_t_ metric on the basis of the input data described below [31]. RIVM’s R_t_ metric is a daily estimate that is based on positive COVID-19 tests in the Netherlands in hospitals from national intensive care foundation (NICE) and from RIVM’s own institutes in municipalities (GGD). When the first symptomatic day of a COVID-19 infected person is not known, RIVM estimates this date. Further, RIVM assumes an average 4 days delay period between infection and first symptoms, and estimate the mean incubation period to be 6.4 days (95% confidence interval (CI): 5.6-7.7) [30].

#### 2.1.2 Meteorological data

Regarding meteorological data, we used datasets from the Royal Dutch Meteorological Institute [32] from February 17, 2020 till September 21, 2020. The downloaded daily data included global solar radiation in J/cm^2^, mean relative atmospheric humidity (% RH), and average temperature in degrees Celsius. For comparison, and given their effects on pollen distribution, we also added precipitation duration in 0.1 hour, precipitation amount in 0.1 mm, mean wind speed, minimum and maximum temperatures in degrees Celsius, mean dew point temperature in degrees Celsius, and sunshine duration in 0.1 hour. Additionally, we calculated the wind chill temperature per day. These datasets were obtained from the KNMI’s centrally located De Bilt weather station. De Bilt is traditionally chosen as it provides an approximation of modal meteorological parameters in the Netherlands, which is a small country. Furthermore, all major population centers in the Netherlands, which account for around 70% of the total Dutch population, are within a radius of only 60 kilometers from De Bilt. We therefore assumed in this study that the measurements from De Bilt are sufficiently representative for the meteorological conditions typically experienced by the Dutch population.

### 2.1.3 Mobility data

We used Google mobility data for relative trends regarding visits to different types of locations in the Netherlands [33] for the same period from February 17, 2020 till September 21, 2020. These location types are: Residential, Workplaces, Indoor Recreation (called retail & recreation by Google, which includes restaurants, cafes, retail, shopping centers, theme parks, museums, libraries, and movie theaters), Outdoor Recreation (called Parks by Google, and including places such as national parks, public beaches, marinas, dog parks, plazas, and public gardens), and Transit Stations (places such as public transport hubs such as subway, bus, and train stations). For comparison, as these are less affected by lockdowns, we also included mobility trends for grocery & pharmacy (places such as grocery markets, food warehouses, farmers markets, specialty food shops, drug stores, and pharmacies).

#### 2.1.4 Seasonal allergens and allergies

For hay fever (allergic rhinitis) we used the data from Nivel [34], for the same period, about weekly incidence reports at primary medical care level, per 100,000 citizens in the Netherlands. Primary medical care is the day-to-day, first-line healthcare given by local healthcare practitioners to their registered clients as typical for the Netherlands. The hay fever incidence metric is a weekly average based on a representative group of 40 primary care units, and calculated using the number of hay fever reports per primary care unit divided by the number of patients registered at that unit. This is then averaged for all primary care units and then extrapolated to the complete population. We used interpolation to generate a daily data set.

For comparison, we included daily mean pollen concentrations based on the data from two Dutch pollen stations: Elkerliek Ziekenhuis in Helmond (latitude 51.487059, longitude 5.662036) [35], and Leiden University Medical Center in Leiden (latitude 52.166309, longitude 4.477315) [36]. The mean pollen concentration is measured in grains/m^3^, whereby we used the daily totals for the 42 types of pollen particles for which by both stations the numbers are counted and averaged per day per 1 m^3^ of air. The common Burkard spore trap is used by these stations. It was noticed before that a metric including all available allergenic particle types, lower allergenic or higher allergenic, correlates stronger with the incidence or R_t_ of COVID-19, than a metric only based on higher allergenic particle types [9, 11].

### 2.2 Data sets consolidation

For all datasets we have selected the same overlapping period of COVID-19 and the first full allergy season [8], during 2020. The overlapping period runs therefore from February 17, 2020 till September 21, 2020 (n = 218 days), when the total pollen concentrations structurally drop below 10 grains/m^3^ as an indication for the end of pollen season.

For sensitivity analyses, we also extended the datasets to periods till June 10, 2021 (n = 480 days).

For mobility datasets the clearly intra-week patterns required a 7 days moving average to reduce noise. Therefore, for reasons of consistency, we calculated 7 days moving averages for all other variables as well.

### 2.3 Statistical analysis

Variables are presented with their sample sizes (n), means (M), and standard deviations (SD). We calculated correlation coefficients to assess the strength and direction of relations of each independent variable with R_t_, and with each other.

Stepwise backward multiple linear regression for all independent variables on R_t_ was used to keep only candidate predictors that are significant (p < 0.05) in the model and remove insignificant predictors. Next, we removed predictors that were multicollinear as defined below. With the remaining independent variables the F-value, standard deviations and errors, degrees of freedom (DF), and significance level, are calculated to test the goodness of fit hypothesis for our predictive model for R_t_. Further, the multiple R, Multiple R squared (R^2^) and adjusted R^2^ correlation coefficients are calculated to estimate the predictive power of our model. Additionally, the algebraic equation to predict R_t_ is determined, which is just to be understood as an empirical formula. Per independent variable the (standard) coefficient, standard error, t-stat and its 95% CI, probability, and the variance inflation factor value (VIF) are calculated.

Further, as linear regression assumes normality of the residuals, we applied the Shapiro-Wilk test and to test the homoscedasticity requirement – homogeneity of variance of residuals– the White test is applied. To analyze multicollinearity we used a VIF value of 2.5 as a threshold. Additionally, the priori power is calculated of each predictor alone and compared with the full model. Although the independent variable R_t_ assumes time lags, we also studied the autocorrelations of residuals, whereby we interpret an autocorrelation beyond a time lag of 7 days as an indication that our model probably might miss a key predictor. Finally, we created calibration plots to visually review the fit of the model.

For selected independent variables with a p value < 0.05 and VIF score < 2.5, standard log_10_, square root and quadratic (^2) data transformations are applied to reduce non-linearity in relations between variables which helps to reduce skewness, and, especially, meet the normality and homoscedasticity requirement. Such data transformations do not change the nature and direction of relations between independent variables and R_t_. In case of the relative mobility trend data we added a constant before such data transformations to avoid loss of data because of negative numbers. For other variables that was not necessary as they only included positive numbers.

We reported the results in APA style, adapted to journal requirements, and applied the TRIPOD guidelines in so far applicable

All statistical analyses were done with Stats Kingdom 2021, which we benchmarked on R version 3.5.

## 3.Results

### 3.1 Variables and their correlations

The sample sizes (N), means, and SDs of the independent variables as used in our multiple linear regression models are summarized in Table 1. The values are given for the data sets after applied data transformations.

**Table 1:**
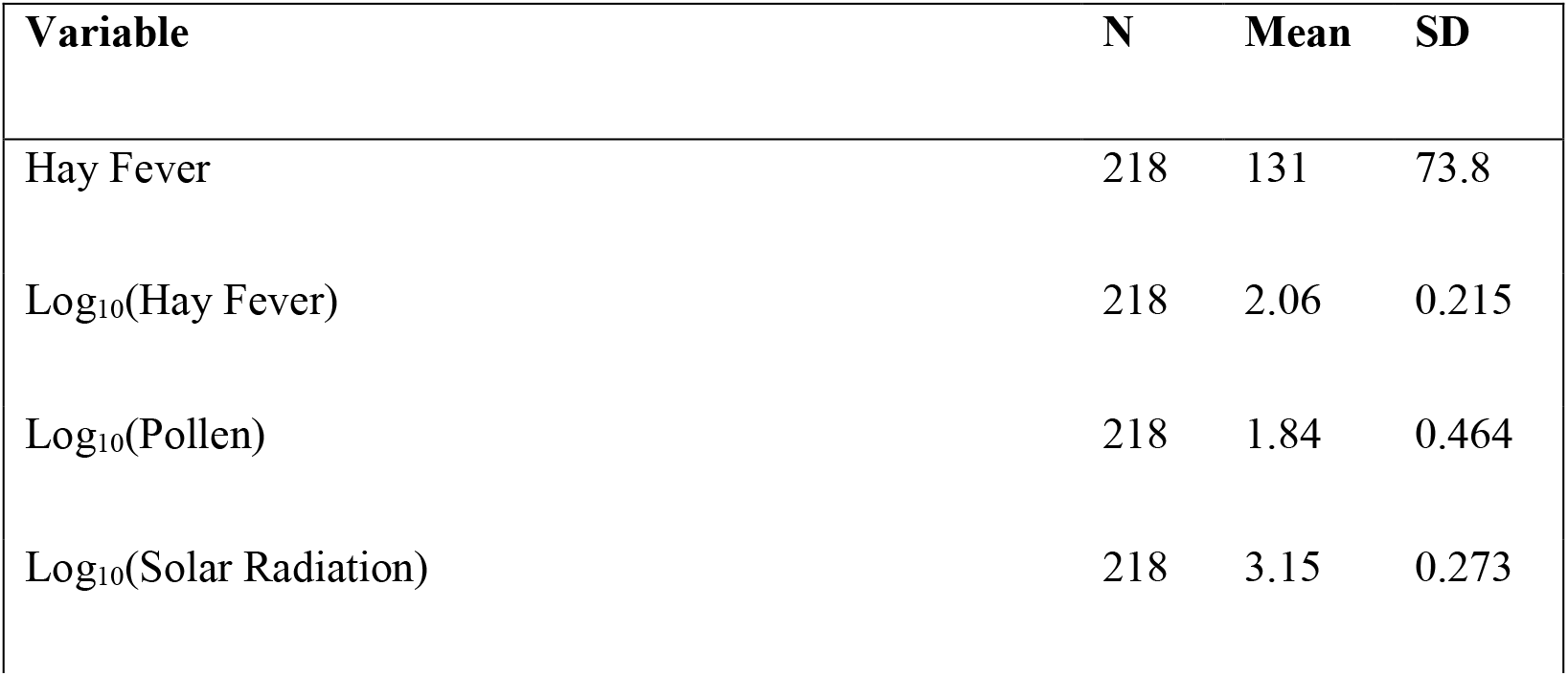
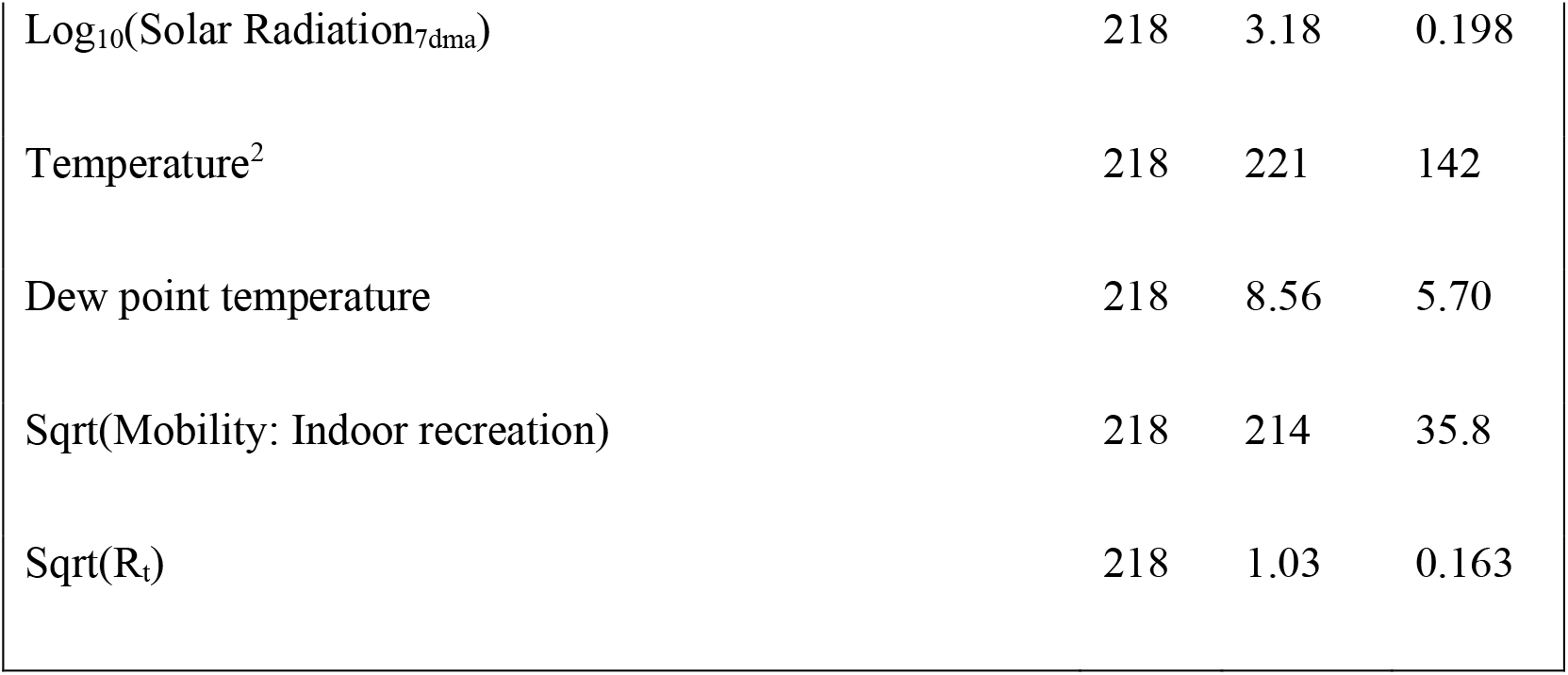
overview means (M), standard deviations (SDs) and skewness values. Overview of mean (M), and standard deviation (SD) per independent variable as used in the multiple linear regression models. The function Sqrt returns the square root of the variable.

During the allergy season, the factors that negatively correlate with R_t_, are in order of strength: hay fever (r(218) = -0.65, p < 0.00001), solar radiation (r(218) = -0.63, p < 0.00001), pollen (r(218) = - 0.62, p < 0.00001), and temperature (r(218) = -0.12, p = 0.085). Positively correlated to R_t_ are relative humidity (r(218) = 0.55, p < 0.00001) and the related dew point temperature (r(218) = 0.12, p = 0.082). Further, higher relative humidity is associated with rain or fog, and thus reduced solar radiation and lower temperature. Temperature and solar radiation are associated as well, although only moderately strong: r(218) = 0.39, p < .00001).

Pollen and hay fever are, as to be expected, associated: r(218) = 0.50, p < .00001), although moderately strong. We did not add allergenicity weights to different pollen particles, and the pollen stations do not cover all types of allergenic particles such as, for example, mold spores. Therefore, having both data sets next to each other has added value, at least for our environmental model. Solar radiation is an important factor as it has, during allergy season, stimulating effects on pollen (r(218) = 0.40, p < .00001) and subsequently hay fever (r(218) = 0.40, p < .00001), in addition to its associations with temperature and R_t_.

The mobility places that are correlated with R_t_ are Indoor Recreation (n(218) = 0.761, p < .00001), Residential (n(218) = -0.684, p < .00001), Transit Stations (r(218) = 0.563, p < .00001), Workplaces (r(218) = 0.532, p < .00001), Grocery & Pharmacy (r(218) = 0.472, p < .00001), and, not significantly, Outdoor Recreation (r(218) = -0.048, p = 0.5). Indoor Recreation and Residential are most strongly inverse correlated: r(218) = -0.817, p < .00001), and thus highly collinear (p > 0.8). Indoor Recreation has moderately strong positive correlations with all other mobility variables, and should therefore seen as a representant of a cluster.

Temperature and dew point temperature had a high correlation of r(218) = 0.84 (p < 0.00001), and appear thus to be collinear. These variables although they have, standalone, no significant correlation with R_t_, still play a role in our combined and environmental model, probably because of their indirect effects on mobility and pollen maturation and dispersion, with their opposite associations with R_t_.

### 3.2 Outcomes combined model

After several iterations with stepwise backward multiple linear regression, four independent variables were selected from the combined pool of environmental and mobility variables that are both significant (p < 0.05) and have a VIF value below 2.5. These selected predictors are: temperature, solar radiation, hay fever, and Indoor Recreation (see Table 2). From the mobility datasets, residential was significant as well but was deselected based on its very high multicollinearity with all other mobility variables, homoscedasticity concerns and lowered explanatory power. In other words, staying at home has a beneficial effect, but, does not explain at which out-of-home location most COVID-19 infections occur. Without the hay fever data, the pollen data would have been significant, but using only the pollen data led to homoscedasticity concerns, which were fully mitigated when using the hay fever data instead.

**Table 2:**
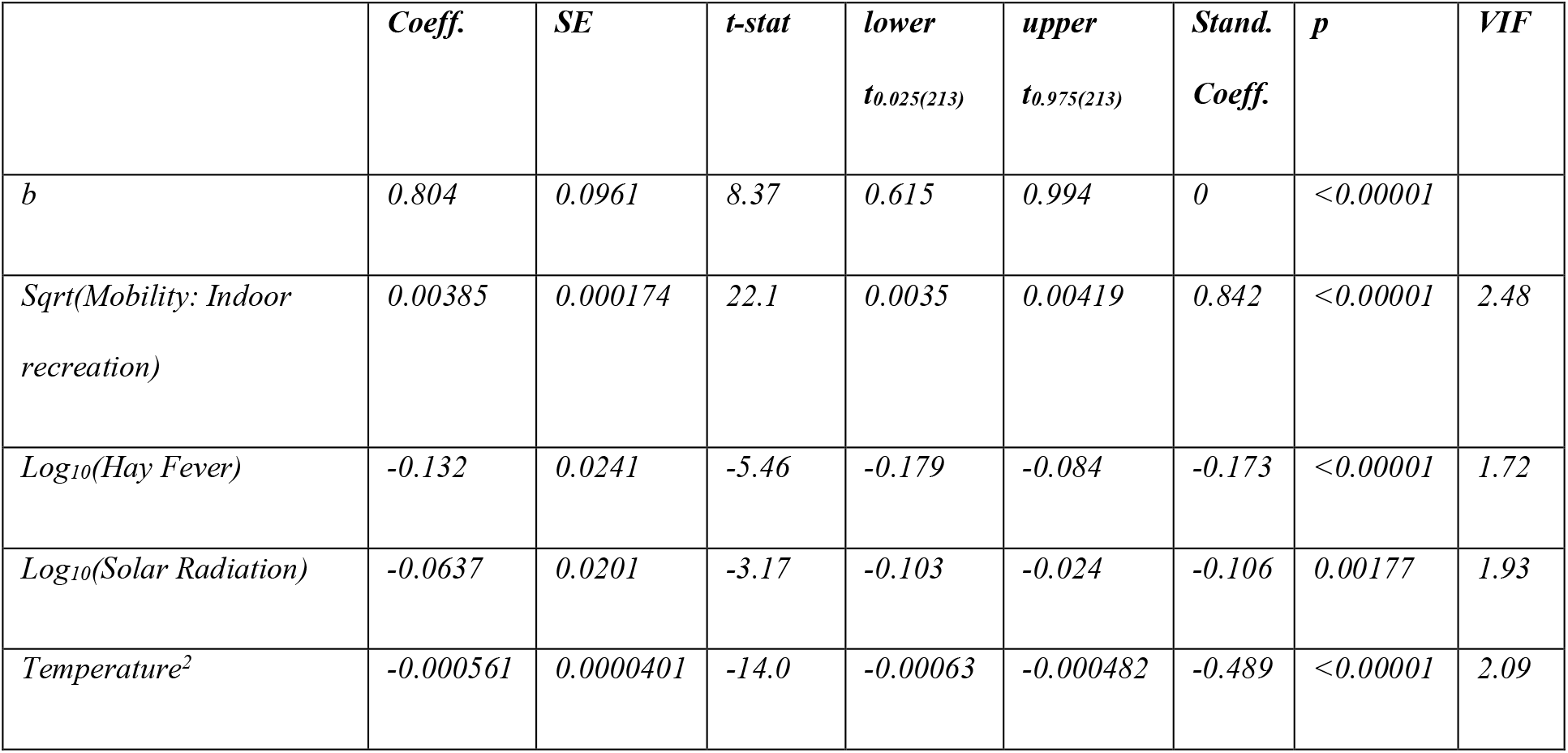
multiple linear regression for mobility *and* environmental predictors. Overview of outcomes per predictor after multiple linear regression for both mobility and environmental variables. Selection of predictors is based on being (highly) significant and having multicollinearity (VIF) score below 2.5. The function Sqrt returns the square root of the variable.

|  | *Coeff.* | *SE* | *t-stat* | *lower*<br>*t<sub>0.025(213)</sub>* | *upper*<br>*t<sub>0.975(213)</sub>* | *Stand.*<br>*Coeff.* | *p* | *VIF* |
| --- | --- | --- | --- | --- | --- | --- | --- | --- |
| *b* | *0.804* | *0.0961* | *8.37* | *0.615* | *0.994* | *0* | *<0.00001* |  |
| *Sqrt(Mobility: Indoor recreation)* | *0.00385* | *0.000174* | *22.1* | *0.0035* | *0.00419* | *0.842* | *<0.00001* | *2.48* |
| *Log<sub>10</sub>(Hay Fever)* | *-0.132* | *0.0241* | *-5.46* | *-0.179* | *-0.084* | *-0.173* | *<0.00001* | *1.72* |
| *Log<sub>10</sub>(Solar Radiation)* | *-0.0637* | *0.0201* | *-3.17* | *-0.103* | *-0.024* | *-0.106* | *0.00177* | *1.93* |
| *Temperature<sup>2</sup>* | *-0.000561* | *0.0000401* | *-14.0* | *-0.00063* | *-0.000482* | *-0.489* | *<0.00001* | *2.09* |

On the basis of the multiple linear regression test, we can reject the null-hypothesis (H_0_) that our combined predictive model with the four selected factors does not provide a good fit: F_(4, 213)_ = 374.2, p < 0.00001. R^2^ equals 0.875, which means that our predictors explain 87.5% of the variance of R_t_. The adjusted R square equals 0.873, and the coefficient of multiple correlation (R) equals 0.936. A simple Pearson correlation between our model’s predicted and the observed values for R_t_ is equally strong and highly significant: r(218) = 0.996, p < .00001. It means that there is a strong, and highly significant, relationship between our combined model’s predicted and the observed R_t_ of COVID-19 (see Fig. 1 and Fig. 2).

**Figure 1:**
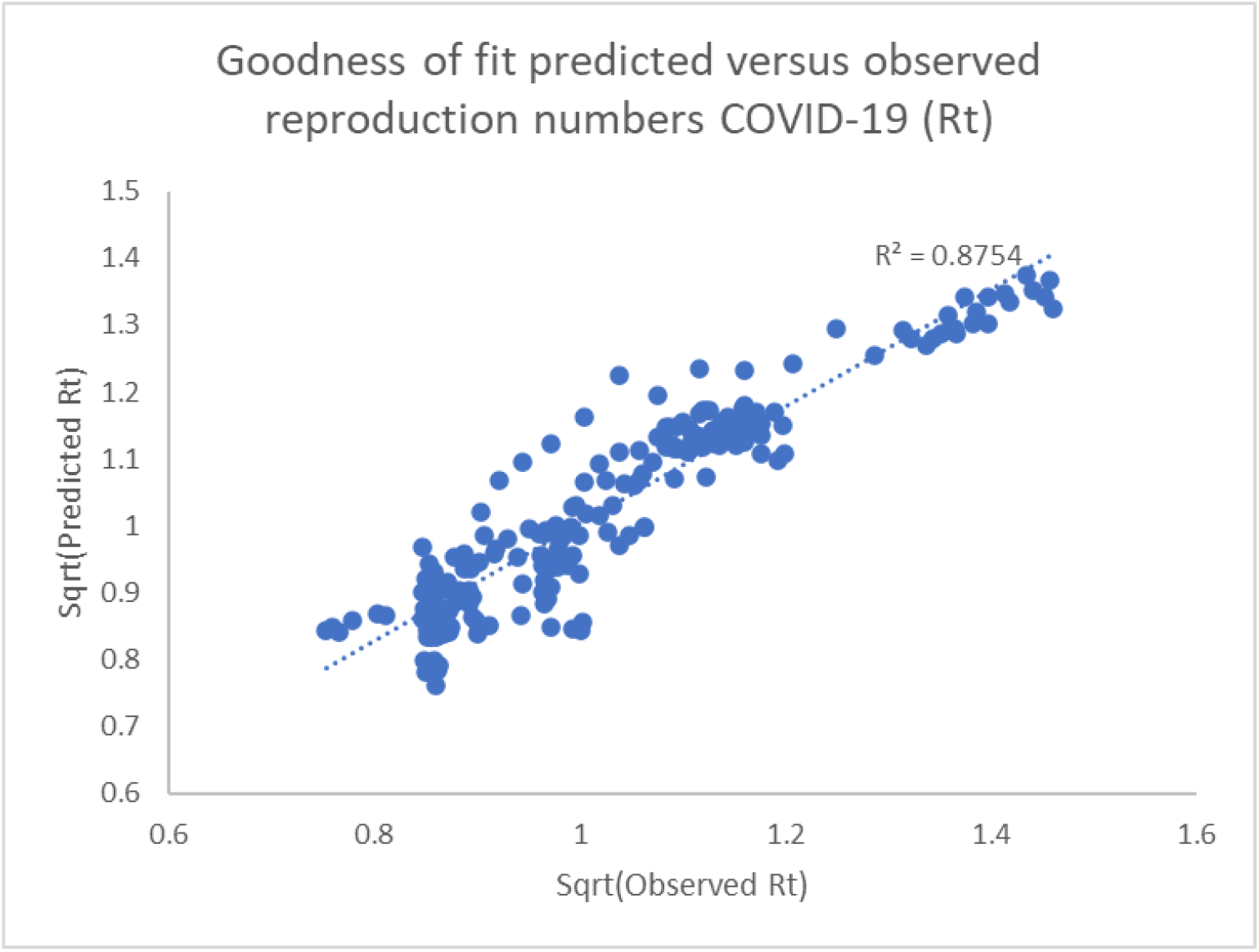
scatter diagram predicted versus observed reproduction number. The combined mobility and environmental model is superior as its predictions 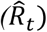 explain 87.5% of the variance of the observed reproduction number of COVID-19 (R_t_) during allergy season.

**Figure 2.**
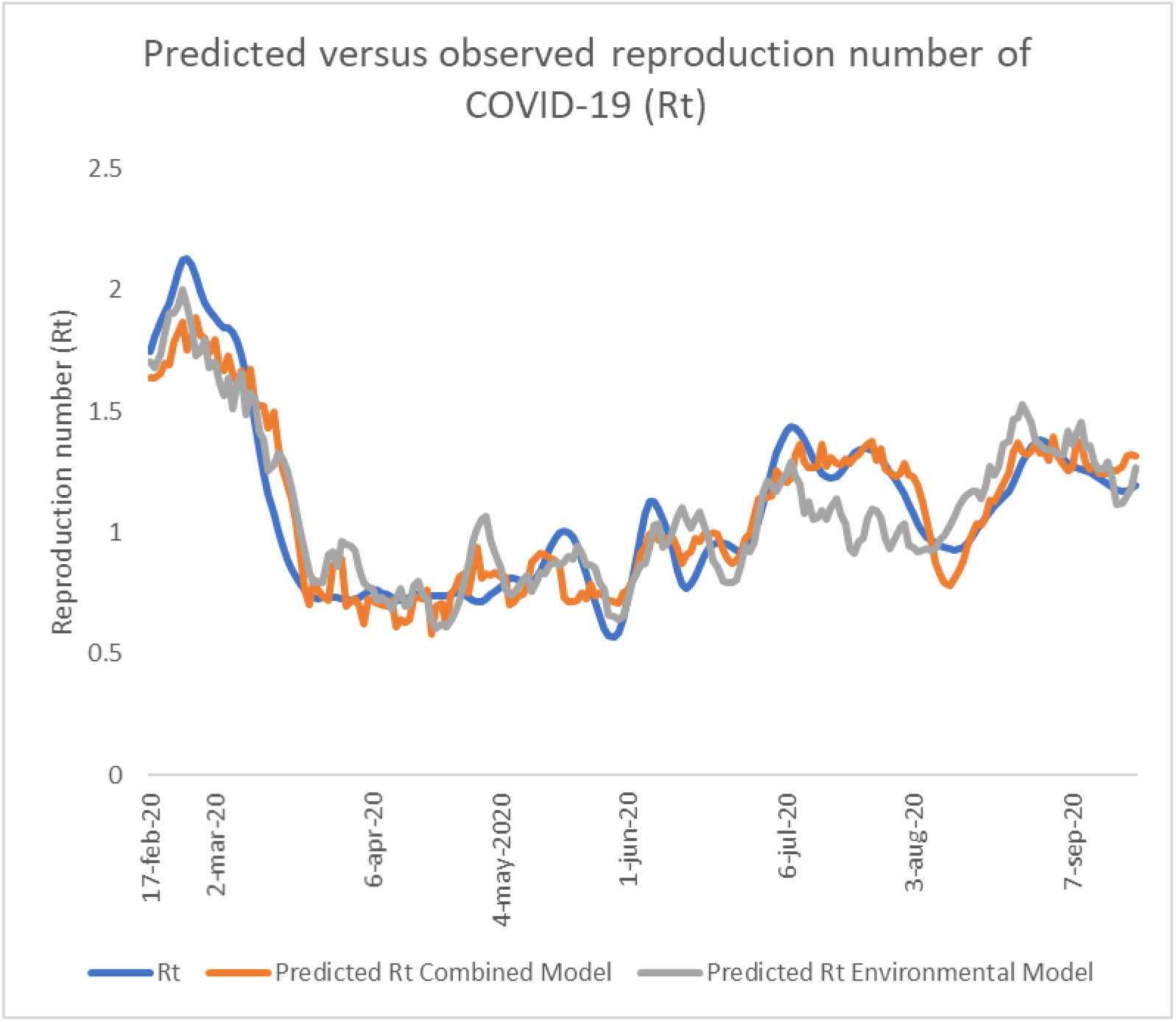
Time series predicted versus observed reproduction number COVID-19. The time series of the predicted 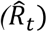 versus the observed reproduction number of COVID-19 (R_t_) in the Netherlands show the very good fit of both the combined and environmental model during allergy season in the Netherlands. However, the Combined Model predicts Rt even better. The seasonality effect in March is visible in both model.

The combined predictive model’s regression formula looks as follows:

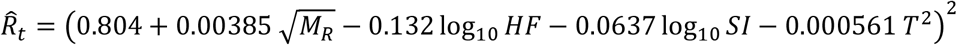

Where 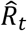 is the predicted effective reproduction number for COVID-19, *M*_*R*_ is the indexed mobility trend data for Indoor Recreation locations to which the mobility constant of 60,000 is added, *HF* is the hay fever incidence per 100K citizens, *SI* is the mean global solar radiation in J/cm^2^, and *T* is the mean temperature in degrees Celsius.. In our dataset, the transformed variables only contain positive numbers.

### 3.3 Statistical outcomes environmental model

For the environmental model we excluded mobility data. Again the solar radiation and hay fever were selected as predictor of R_t_. The pollen metric added explanatory power, and dew point temperature was selected at the expense of its collinear, temperature (see Table 3). Relative humidity was again deselected as an insignificant predictor.

**Table 3:** multiple linear regression for environmental predictors *only*. Overview of outcomes per selected environmental predictor after multiple linear regression. Selection of predictors is based on being (highly) significant and having multicollinearity (VIF) score below 2.5.

|  | Coeff. | SE | t-stat | lower<br>t<sub>0.025(213)</sub> | upper<br>t<sub>0.975(213)</sub> | Stand.<br>Coeff. | p | VIF |
| --- | --- | --- | --- | --- | --- | --- | --- | --- |
| b | 3.00 | 0.100 | 30.0 | 2.80 | 3.19 | 0 | <0.00001 |  |
| Log<sub>10</sub>(Pollen) | -0.0587 | 0.0144 | -4.08 | -0.0870 | -0.0303 | -0.167 | 0.0000633 | 1.56 |
| Log<sub>10</sub>(Solar radiation<br>7dma) | -0.592 | 0.0370 | -16.0 | -0.664 | -0.519 | -0.717 | <0.00001 | 1.89 |
| Dew point temperature | 0.00674 | 0.00109 | 6.19 | 0.00459 | 0.00888 | 0.235 | <0.00001 | 1.35 |
| Hay fever | -0.000262 | 0.0000903 | -2.91 | -0.000440 | -0.0000844 | -0.118 | 0.00405 | 1.56 |

On the basis of the multiple linear regression test, we can reject the H_0_ that our environmental predictive model with the four selected factors does not provide a good fit: F_(4, 213)_ = 181.3, p < 0.00001, and R^2^ equals 0.773, which means that our environmental predictors explain 77.3% of the variance of R_t_. The adjusted R^2^equals 0.769, and the coefficient of multiple correlation (R) equals 0.879. It means that there is a very strong direct and highly significant relation between our environmental model’s predicted and the observed reproduction numbers of COVID-19.

The environmental model’s regression formula looks as follows:

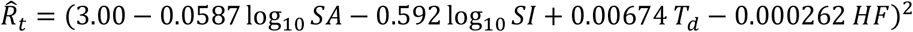

Where R_t_ is the predicted reproduction number for COVID-19, SA is average seasonal allergens or pollen concentrations in particles/m^3^, SI is the 7 days moving average of global solar (ir)radiation in J/cm^2^, T_d_ is the average dew temperature in degrees Celsius, and HF is the hay fever incidence per 100K citizens. In our dataset, the transformed variables only contain positive numbers.

## 4. Discussion

The predictive power of the combined environmental-mobility model including solar radiation, hay fever, temperature and visits to Indoor Recreation locations (87.5%) surpasses the environmental model (77.3%) with more than 10%. Furthermore, the improved accuracy of the combined model shows that adding mobility trends not only helps to control the environmental model for lockdown effects, but also clearly improves it by helping to show the importance of seasonal behavior better. For example, nice weather (sun shine, warm, low humidity) in The Netherlands is related to higher pollen concentrations, and more visits to crowded non-residential locations where social distancing is hard to apply. The latter is in turn associated with increased COVID-19 infections. Interestingly, increased visits to Outdoor Recreation locations are not associated with an increase in COVID-19 infections (R_t_). This finding suggests that outdoor transmission of SARS-CoV-2 is far less likely than indoor transmission, and that restrictive policies that limit visiting Outdoor Recreation locations have less added value.

Although, overall, the environmental model is weaker than the combined model, it is still somewhat better at the onset of COVID-19 during February and March 2020. This is probably explained by the exclusion of ski holiday locations abroad, in Italy and Austria, where many of the first patients contracted COVID-19, which leads to an underestimation of both the Indoor Recreation and Outdoor Recreation trend. On the other hand, the combined model is somewhat better in July when lockdown restrictions were relaxed and people were less strict, which is caught well by the mobility trends variable. Both models are almost equally strong in predicting the seasonal decline in March/April, which indicates that the relative importance of restrictive measures was probably not the main driver of that particular decline, but the seasonality effect was.

Of the non-residential locations, especially Indoor Recreation is by far the best predictor of increasing COVID-19 infections (R_t_), which makes sense as social distancing in busy shopping locations, bars, discos and other such locations, is hard to maintain. Especially, when the seasonality effects are offset by relaxed lockdown measures and social distancing discipline. Even more, if people are under the influence of alcohol and party drugs in crowded party locations, social distancing becomes a distant reality. Additionally, the strong inverse correlation of Residential with R_t_, shows that staying at home, because of lockdown measures, is effective. That all other indoor locations have a positive correlation with R_t_, shows basically the same: when lockdown measures are relaxed, infection rates increase as people will meet more other people.

The single effect of high temperature on R_t_ appears to be not significant. The role of temperature can be understood only when its associations with other variables such as mobility trends and pollen maturation and dispersion are taken into account. Humidity in general, relative or specific (T_d_), appears to be positively associated to COVID-19 reproduction, as it is associated with a reduced solar radiation and seasonal allergens, and more traffic to indoor locations which are associated with an increase in infections. Even despite observations that, indoors, very dry air, with a low absolute humidity, might favor SARS-CoV-2 transmission, which is likely caused by increased aerosolisation of infectious aqueous particles. Finally, although we assume that day length is associated to solar radiation, it might still be interesting to look if this solar-related variable could add something to the predictive power of our models.

### Methodological concerns

Test bias, especially for new viruses such as COVID-19, is a major methodological challenge. The approach to use more reliable metrics such as the number of hospitalizations to generate the R_t_ metric appears to be a good method to reduce test bias. But, the change of methodology in June 2020, when more test stations were included with their fluctuating test capacities, most likely led to the introduction of test bias in the R_t_ metric. Such reliability concerns may have reduced the predictive power of our combined and environmental model.

The usefulness of the pollen concentration metric might be improved by taking into account the allergenicity per particle type. The allergenicity classification is available, but it is not on a ratio scale and there are discussions about the accuracy of this classification. Furthermore, other allergenic particles like mold spores, are hardly ever covered by European pollen stations because of budget constraints.

We observed that the Indoor Recreation and Outdoor Recreation metric might need to be expanded to holiday locations in foreign countries. Unfortunately, that is something that is currently not possible via the Google Mobility datasets.

In our research we precluded the period of intensive vaccination from January 2021 onward. On the other hand, if the protective immunity would be short-lasting, we might still be confronted with resurgences of COVID-19 [37], and it would be of interest to test the predictive models for such events. It is likely that new waves, will be less intense and short-lived given longer lasting B-cell and T-cell memory of people that have been infected or are vaccinated already. Therefore, it might be good to control for herd immunity levels when testing the predictive models for subsequent allergy seasons.

Additionally, it might be of interest to differentiate R_t_ per virus variant, given that genetic drift typically leads to more contagious but less deadly variants, that somewhat change the dynamics of COVID-19.

Finally, testing the predictive models for a wider geographical scope would be of interest, but would require metrics that are not widely available such as a standardized metrics for R_t_, hay fever incidence, and pollen datasets.

## Conclusion

The combined, mobility and environmental, model explains 87.5% of the variance of R_t_ of COVID-19 during spring season in a country in the temperate climate zone like the Netherlands, and provides a very good fit (F_(4, 213)_ = 374.2, p < 0.00001), as the predicted and observed R_t_ correlate strongly and highly significantly. The significant predictors in the combined model are temperature, solar radiation, hay fever incidence, and the Indoor Recreation trend. The environmental factors are inversely associated with R_t_, On the other hand, more visits to Indoor Recreation locations is associated with more infections (R_t_). This seems to be the best mobility predictor for the effects of lockdown measures on the spread (R_t_) of COVID-19. On the other side of the spectrum, moving to Outdoor Recreation locations is not significantly associated with changes in R_t_, and including such locations in lockdown regimes appears to be ineffective.

The solely environmental model, is around 10% less powerful than the combined model. Nevertheless, the environmental model shows that pollen concentrations and dew point temperature as a collinear of temperature, have an added explanatory value. Further, there are short periods in which the environmental model beats the combined model.

## Data Availability

We included links to the (open/public) data sources used in our manuscript under references.

## ACKNOWLEDGEMENTS

Thanks to Geert H. Keetels, Bas J.H. van de Wiel of Delft University of Technology for help with dew point temperature calculation, and discussing insights related to specific humidity and COVID-19.

Further, thanks to Letty de Weger of Leiden Univsity Medical Center for providing pollen statistics and providing useful insights.

## FOOTNOTES PAGE

**Martijn Hoogeveen:** Conceptualization, Methodology, Data Curation, Formal Analysis, Writing, Investigation, Visualization, Resources.

**Ellen Hoogeveen:** Methodology, Writing, Resources.

**Aloys Kroes:** Conceptualization, Reviewing, Data Curation.

### Data statement

The links to downloadable datasets are provided in the reference list or references to the sources are included. Upon request, the data used for this manuscript is available for inspection, but for other purposes we kindly refer to the respective copyright-holder(s).

